# Skin cancer high-risk patient screening from dermoscopic images via Artificial Intelligence: an online study

**DOI:** 10.1101/2021.02.04.21251132

**Authors:** David Coronado-Gutiérrez, Carlos López, Xavier P. Burgos-Artizzu

**Affiliations:** Transmural Biotech S. L. Barcelona, Spain

**Keywords:** Dermatology, Dermoscopy, Skin Cancer, Deep Learning, Artificial Intelligence

## Abstract

**Objectives:** To evaluate a novel Artificial Intelligence (AI) method for the detection of malignant skin lesions from dermoscopic images.

**Methods:** 58,457 dermoscopic images available online from the International Skin Imaging Collaboration (ISIC) Archive were downloaded. These images were acquired from different centers worldwide by recognized dermatologists and show varied clinical outcomes belonging to different types of benign and malign skin lesions. A state-of-the-art AI skin lesion classifier based on Deep Learning was designed. The method, fully automated, first locates and segments the nevus in the image and then classifies it into either benign or malign type.

**Results:** 1,631 images (2.8%) were discarded due to bad quality. A total of 56,826 images were finally used. Two thirds of the images (37,688) were used to train the classifier, leaving the remaining 19,138 images for validation. In this set, malignant lesions had a prevalence of 15.4% (2,956/19,138). The AI skin lesion classifier reached an area under the curve (AUC) of 87.4%. Optimal cut-off point in terms of accuracy resulted in an 85.9% accuracy (16,439/19,138) and sensibility of 89.6% (2,648/ 2,956) at 85.2% (13,791/16,182) specificity. Negative predictive value (NPV) was 97.8% (13,791/14,099). Other training/validation splits were also evaluated, showing similar results.

**Conclusions:** A novel AI method showed promising results as skin lesion classifier from dermoscopic images. Its high NPV value could make it suited for high-risk patient screening. A large clinical study to confirm these results is needed and will be pursued.

## INTRODUCTION

Skin cancer is the most common cancer worldwide in white populations^1^. This type of cancer shows an increasing incidence rate but a stable or decreasing mortality rate^2^. Only in the United States, this type of cancer causes 5 million cases annually and the annual cost of care exceeds $8 billion^3,4^. Melanoma, the most dangerous type, leads to over 9,000 deaths per year, especially because of its delayed diagnosis^5^. Early detection makes a difference, 5-year survival rate for patients in the U.S. whose melanoma is detected early. The survival rate drops to 65% if the disease reaches the lymph nodes and 25% if it spreads to distant organs^6^.

The incidence of melanoma in global population varies widely by country, but it is generally very small (less than 1‰ habitants)^2^. For that reason, the general screening for melanoma detection it is commonly performed on people with risk factors such as: high nevus count, high median count of atypical nevi, previous melanoma or family history of melanoma in first -or second-degree relatives^7^. There is no compelling evidence for monitoring low-risk patient groups^8^.

Dermoscopy has had a fundamental role in the early diagnosis of skin cancer and has demonstrated improvement for diagnosis compared to unaided visual inspection^9^. However, clinicians should receive proper training in order to improve the 60% accuracy of unaided inspection to 75-84% of the diagnostic with dermoscopy^9,10^. Several recognition protocols as the ABCD(E) rule or the “ugly duck” sign are used in dermoscopy to improve the detection of melanomas^6,11^. Other recent techniques as Sequential Digital Dermatoscopic Imaging (SDDI) usually combined with Total-Body Photography (TBP), have improved the early detection and could reduce the number of necessary excisions^8,12^.

During the past few years, research has focused on automatic algorithms to improve the current clinical diagnosis from dermoscopic images. Several entities have published large-scale publicly accessible datasets to support researchers in development of automated algorithms^13–15^. The International Skin Imaging Collaboration (ISIC) has developed the ISIC Archive, an international repository of dermoscopic images, for both the purposes of clinical training, and for supporting technical research toward automated algorithmic analysis by hosting the ISIC Challenges.

The rise of artificial intelligence techniques and especially Deep Learning^16^, has increased the number of studies which use this type of algorithms for clinical diagnostic. Several recent studies compared the performance of Convolutional Neural Networks (CNN)^17^ classifiers with clinicians’ in specific binary tasks, such as differentiating malign melanomas from benign nevus^18,19^. Others use CNN to classify multiple types of skin lesions useful for example to be able to differentiate more incident lesions (such as basal cell carcinoma) from other more lethal (such as melanoma)^20–22^. However, none of these studies has been focused on evaluating these algorithms in a broad malign/benign screening scenario.

The main goal of this study was to assess the potential of current state-of-the-art Artificial Intelligence (AI) methods to build a clinical tool able to distinct benign skin lesions from malign ones in a screening scenario of patients with risk factors. To do that, ISIC 2019^23^ and ISIC 2020^24^ datasets were used together to create a large database with a malignant prevalence close to what found in some dermatologic centers during high-risk patient screening.

## MATERIALS AND METHODS

### Study design

ISIC 2019 and ISIC 2020 databases used in this study are composed of dermoscopic images provided from HAM10000 dataset^14^, MSK dataset^13^, BCN_20000 dataset^15^ and ISIC 2020 Challenge dataset^24^. The images were acquired from different centers, by different operators, using a variety of devices and stored in several formats. The ISIC consortium processed all images, screening them for privacy and quality, and published the images in JPG format. These databases are licensed under a Creative Commons Attribution-NonCommercial 4.0 International License (CC-BY-NC).

The public database of ISIC 2019 is composed by 25,331 training images while the ISIC 2020 contains 33,126 images, for a total when combined of 58,457 images. All images were stored in JPG format with different sizes and resolutions varying from 450×576 pixels to 6000×4000 pixels. All images focus on a main nevus (the one for which a prediction should be made) centered or not. Some images have a ruler, color patch or other artifacts outside the nevus. Figure 1 shows some image examples. Since images are in JPG format, there is no pixel resolution information and therefore there is no information about the real size of the lesions which could be useful to train the classifier.

**Figure 1.**
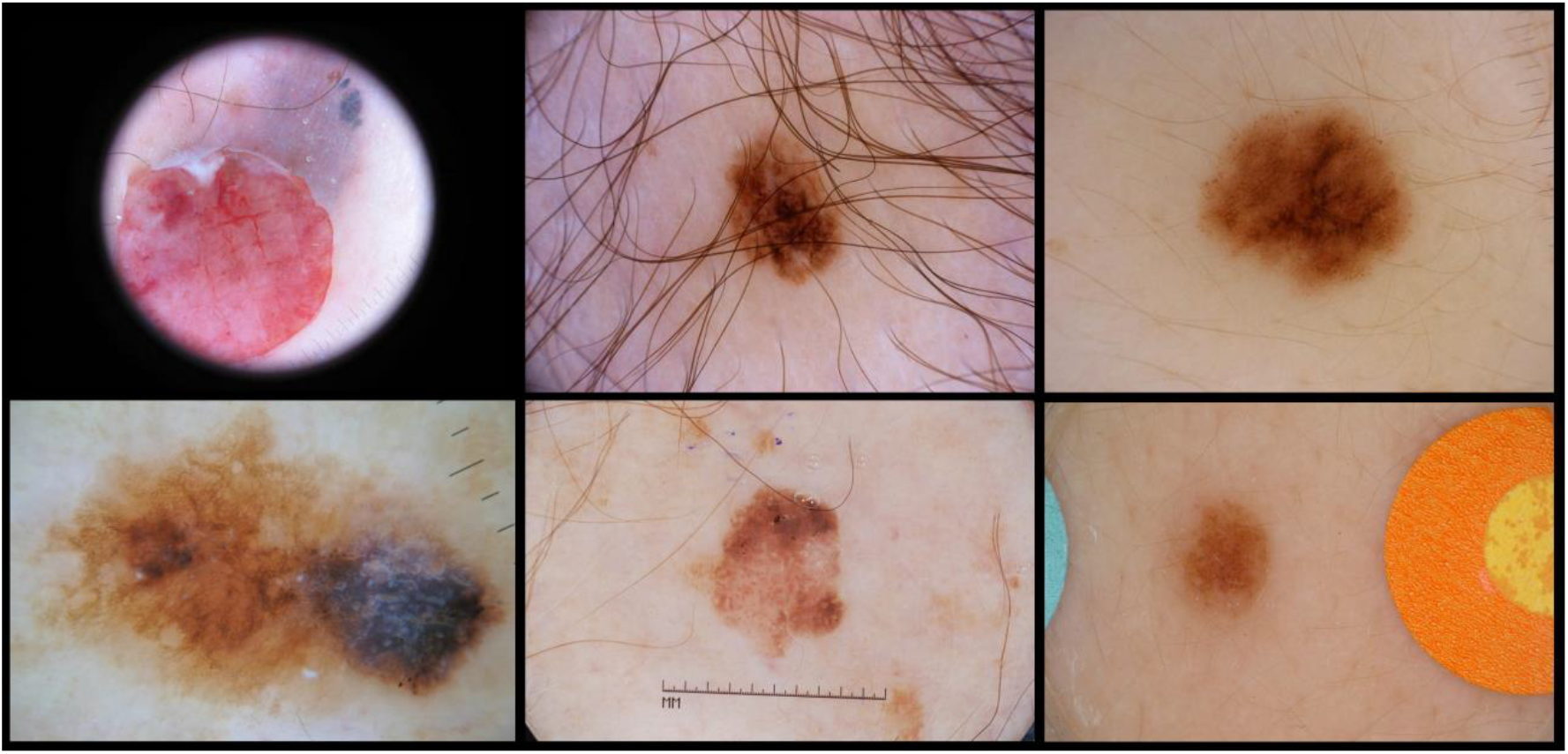
Image examples from ISIC database.

All images have associated skin lesion type, vetted by recognized dermatologists. Images contain different types of skin lesions, some of them benignant: melanocytic nevus, actinic keratosis, benign keratosis, dermatofibroma, vascular lesion and lentigo NOS; and others malignant: melanoma, basal cell carcinoma and squamous cell carcinoma. Also, most of images have associated information about the lesion location in the body and the patient’s age and gender.

### Automated AI skin lesion classifier

A novel method for skin lesion classification from dermoscopic images was developed using state-of-the-art Deep Learning techniques. We coined this new method quantusSKIN. It is formed by two distinct modules: the first locates and segments the nevus in the image and the second one classifies it into belonging either to a benign or malign skin lesion.

#### Nevus detection and segmentation

Given that images have different resolutions, the nevus is not always centered and other elements can be present in the image (color patches, secondary nevus, rulers or other artifacts). A segmentation algorithm was developed to automatically delineate the main nevus’ region of interest (ROI) from each image. To build the algorithm, a set of 2,877 images from MSK dataset^13^ were used. These images contained the manual delineation (provided by clinicians) of the nevus’ region.

A fully convolutional network (FCN8s)^25^ was used to do the segmentation task. The network was trained using Transmural Biotech’s AI platform^26^. Of the 2,887 images, 2,114 were used to train the network and the 773 remaining were used to test the algorithm and obtain the performance of the segmentation task.

#### Skin lesion classifier

Once the main nevus detected and segmented, a CNN^17^ classifier was adapted for the task. More specifically, a variant of the Inception algorithm^27^ was used. The network of the prediction algorithm was trained using Transmural Biotech’s AI platform^26^. The classifier was trained using softmax cross-entropy loss and adam optimizer. 10% of the training set was used as validation set. The first training using ImageNet images was performed for 90 epochs with a 5 epoch warm-up. Then, the CNN was trained again on training data for a maximum of 30 epochs, early stopping if loss on validation set was not improved for 10 consecutive epochs. Learning rate was adjusted using a cosine decay starting at 1e-4. Batch size was 64 and weight decay was 0.9. To improve learning, data augmentation was used during training. At each batch, images were randomly flipped, cropped between 0 – 10%, translated from 0 – 10 pixels and rotated between [-90; 90] degrees.

### Statistical analysis

Missing information on clinical variables (less than 1% in age and gender and 5% in lesion location) was first tested using Little’s test to verify that blanks were randomly distributed, and then they were filled using multiple imputation. Then, null hypothesis testing was performed to establish the relationship between the clinical outcome (skin lesion type) and the variables, computing significance p-values. More specifically, fisher’s exact test was used for discrete variables (gender and lesion location) while t-test was used for age (normally distributed).

In order to evaluate the accuracy of the segmentation algorithm, each automatic delineation was compared with the manual delineation provided using Jaccard Index. This metric measures similarity between finite sample sets (pixels) and it is defined as the size of the intersection divided by the size of the union of the sample sets.

Finally, to evaluate the skin lesion classifier’s performance, the patients were randomly split into independent training and testing sets, preserving the prevalence of each class type (a.k.a. stratified splits) in each set. Different splits, varying the proportion of training and testing images were benchmarked for completeness. The test set was used to compute output scores for each image and compared with the clinical outcome to obtain the receiver operator characteristic (ROC) curve and compute full area under the curve (AUC). Then, typical statistical metrics (sensitivity, specificity, etc.) were computed from the optimal cut-off point (that maximizing accuracy). For all metrics, 95% confidence intervals were estimated using a 10-fold bootstrap on the test set.

## RESULTS

### Automatic AI nevus segmentation

The automatic delineation on the 773 testing images reached an average Jaccard index of 84.9%, with 14.9% of the ROIs falling below 50% overlap. Figure 2 shows some visual examples of the lesions and their automatic and manual segmentation masks.

**Figure 2.**
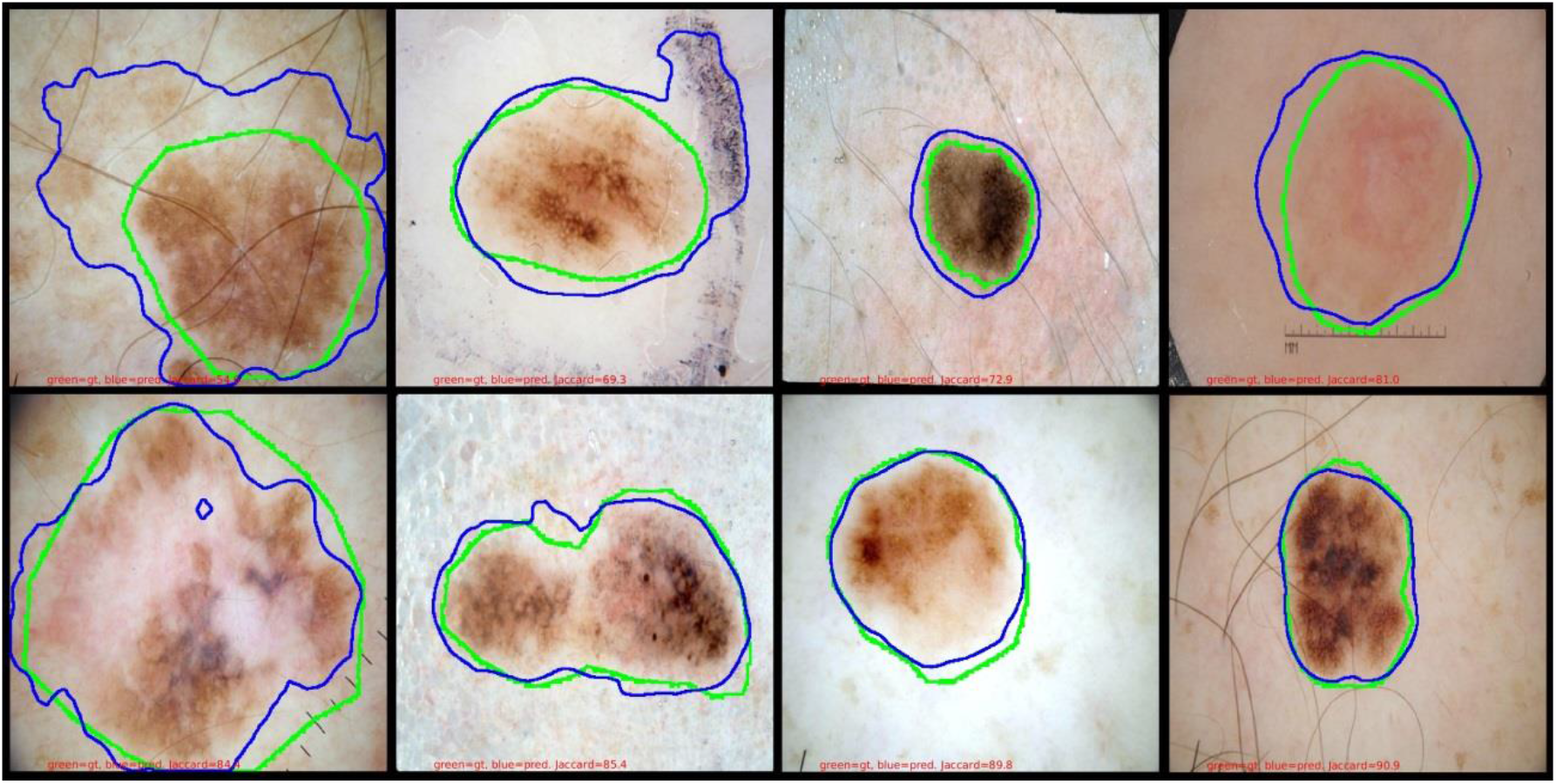
Examples of 8 different lesions and their segmentation masks (green = manual mask; blue = predicted mask).

### Final data used for the study

All 58,457 images from ISIC 2019 and ISIC 2020 were automatically segmented using the created segmenter, obtaining for each image the ROI containing the nevus. 2.8% of the images (1,631) were discarded because the algorithm could not properly delineate the ROIs due to the small size of the lesion or the bad quality of the image. The final database used for the classification task was therefore of 56,826 images.

Of these 56,826 images, 47,953 images contained a benign lesion, while 8,873 images contained a malign one, representing a global prevalence of malignant lesions of 15.6%. This prevalence was in accordance with reported prevalence in high-risk patient screening studies (e.g. 13.6% in *Haenssle et al. 2016*^7^).

Table 1 shows the characteristics of the data population (patient gender, patient age and lesion location in the body), together with null hypothesis testing significance values. Gender and age were significantly related with clinical outcome, with older males having a higher chance of developing a malignant skin lesion. Furthermore, prevalences of malignant lesions varied significantly depending on the skin lesion’ location in the patient body.

**Table 1.** Population characteristics and statistical relationship between variables and clinical outcome.

| Variable | Total | Benignant lesions | Malignant lesions | p-value |
| --- | --- | --- | --- | --- |
| **Age** |  |  |  |  |
| mean (± std) | 51.2 (± 16.2) | 49.0 (± 15.4) | 63.1 (± 15.4) | **0.00** |
| **Gender (%)** |  |  |  |  |
| Male | 29,740 (52.3%) | 24,553/47,953 (51.2%) | 5,187/8,873 (58.5%) | **0.00** |
| Female | 27,086 (47.7%) | 23,400/47,953 (48.8%) | 3,686/8,873 (41.5%) | **0.00** |
| **Lesion Location (%)** |  |  |  |  |
| Head / Neck | 6,426 (11.3%) | 4,223/47,953 (8.8%) | 2,203/8,873 (24.8%) | **0.00** |
| Oral / Genital | 178 (0.3%) | 155/47,953 (0.3%) | 23/8,873 (0.3%) | 0.35 |
| Palms / Soles | 789 (1.4%) | 570/47,953 (1.2%) | 219/8,873 (2.5%) | **0.00** |
| Torso | 27,520 (48.4%) | 24,005/47,953 (50.1%) | 3,515/8,873 (39.6%) | **0.00** |
| Lower extremity | 13,837 (24.3%) | 12,239/47,953 (25.5%) | 1,598/8,873 (18.0%) | **0.00** |
| Upper extremity | 8,076 (14.2%) | 6,761/47,953 (14.1%) | 1,315/8,873 (14.8%) | 0.08 |

### Automatic AI nevus classification

The database was divided into 37,688 images for training the classifier (66% of the total) and 19,138 for testing (33%). Figure 3 shows the ROC curve obtained by the proposed Deep Learning algorithm on the 19,138 testing images. The classifier reached an AUC of 87.4% ±0.1%.

**Figure 3.**
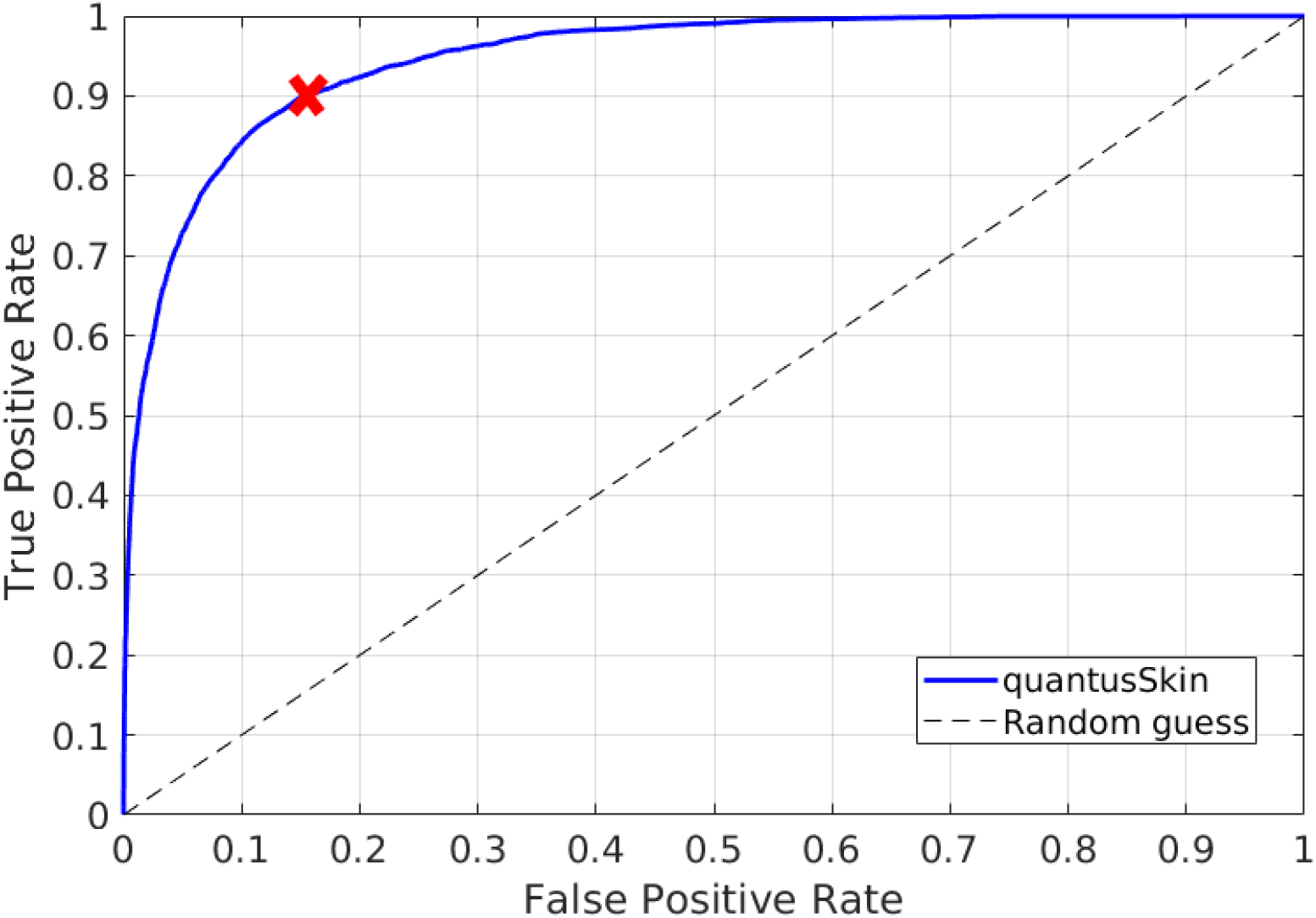
ROC curve for prediction task on the 19,138 test images (66%/33% training-testing split). The red X marks the optimum cut-off point in terms of Accuracy.

Table 2 shows the detailed metric scores at the optimal cut-off point in terms of accuracy computed from the ROC curve. The CNN reached an accuracy of 85.9% with 89.6% and 85.2% of sensibility and specificity, respectively. This scenario would have implied that of the 2,956 patients which had a malignant lesion, the CNN would have successfully detected 2,648, leaving 308 (10%) undetected. Also, of the 16,182 patients with benign lesions, 2,391 (14.8%) would have been falsely flagged. On the other hand, 97.8% (13,791) of the negative classified nevus (14,099) were correctly detected, leaving only 2.2% (308) of false negatives.

**Table 2.**
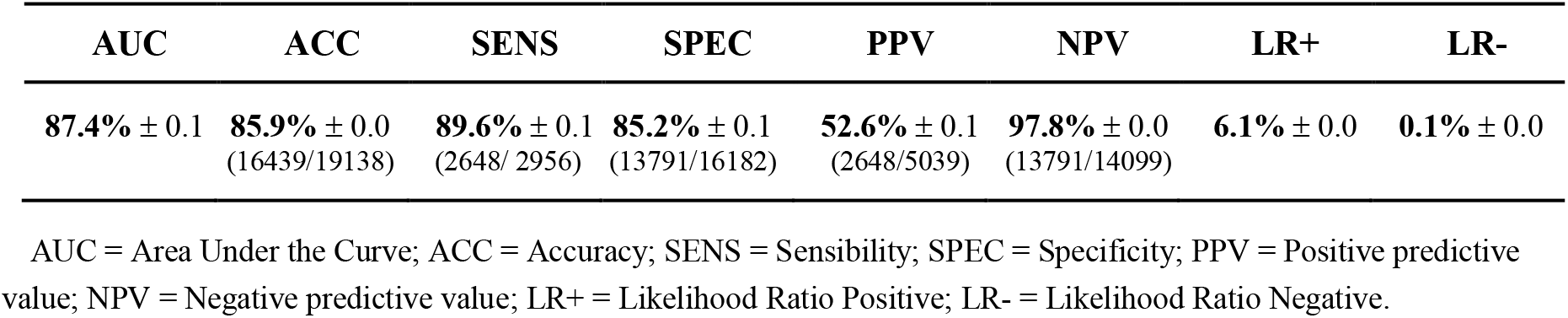
Metric scores at the optimal cut-off point computed from the ROC curve.

Figure 4 shows examples of skin lesion classification on the test images. The first row shows benignant lesions, and the second row shows malignant lesions. The 3 first images of each row correspond to correct classification (true negative or true positive) while the last one shows a failure (false positive or false negative).

**Figure 4.**
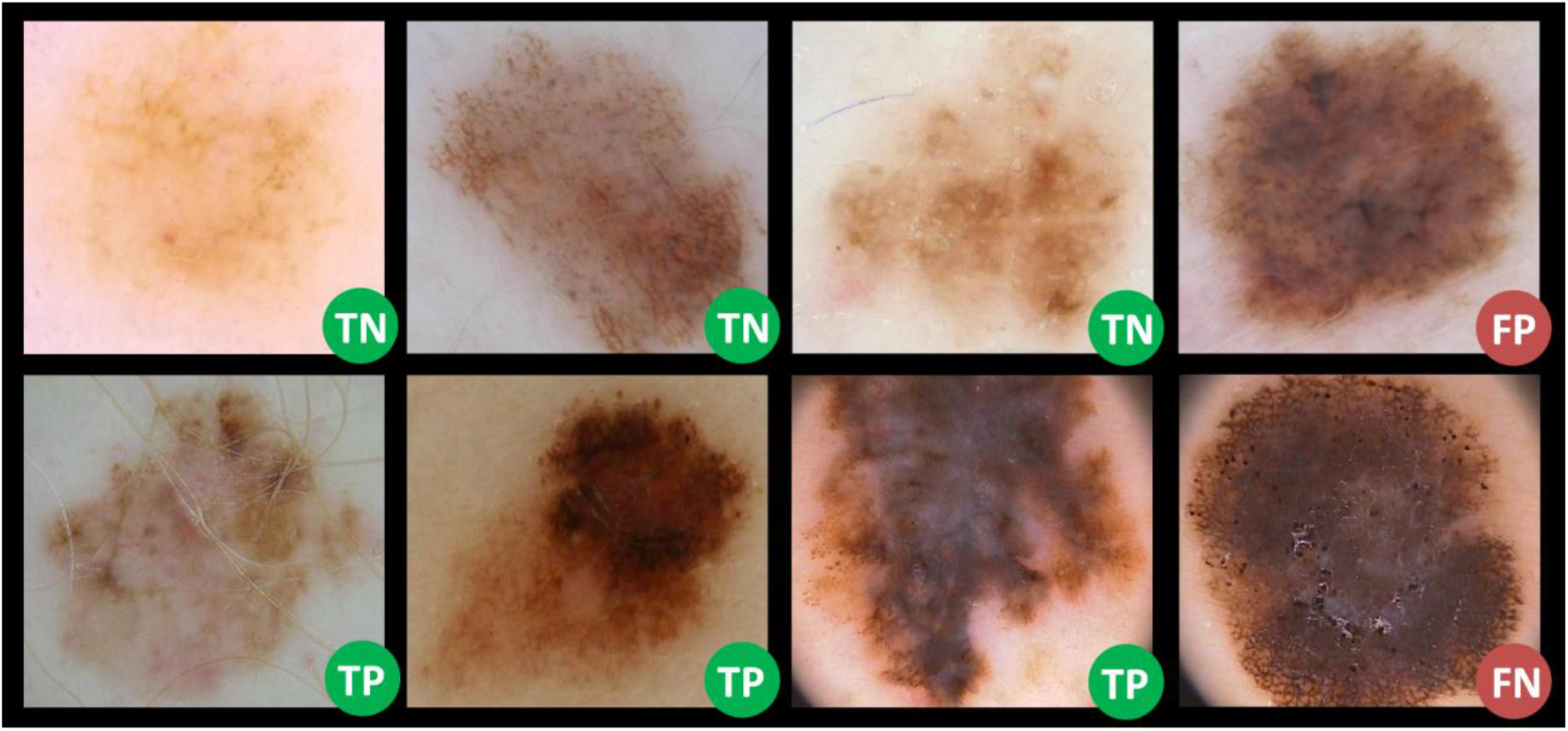
Example of benign lesions (first row) and malign lesions (second row). TN = True Negative; FP = False Positive; TP = True Positive; FN = False Negative.

Finally, figure 5 shows the method’s performance (accuracy and AUC) when other random train-test splits percentages (50/50% and 90/10%) were used compared with the 66/33% split.

**Figure 5.**
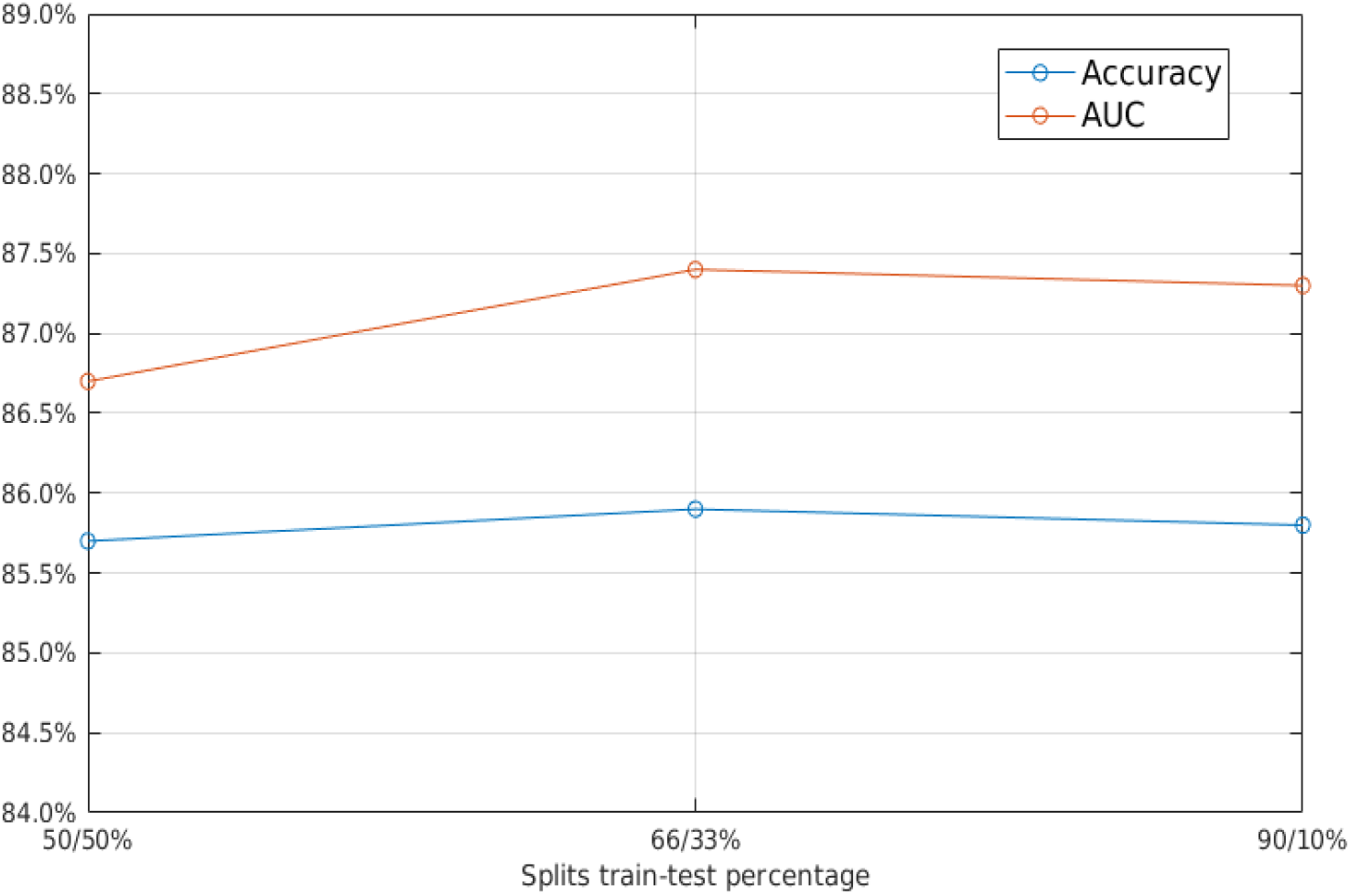
Area Under the Curve (AUC) (in red) and accuracy (in blue) obtained by the classifier when different training/testing splits were used (50%/50%, 66%/33%, 90%/10%).

## DISCUSSION

### Main findings

A novel AI method, based on state-of-the-art Deep Learning, was designed. The novel method is capable of automatically detecting a nevus from a dermoscopic image and then classifies it as benignant or malignant type. It was evaluated on a large set of images downloaded online, showing promising results which open the door to its potential use as screening tool on patients with skin cancer risk factors.

Evaluated on 19,138 images with a high-risk screening realistic malignant incidence of 15.6%, the method reached 85.9% accuracy with a sensibility of 89.6% at 85.2% specificity. Its negative predictive value was very high: 97.8%. In addition, the method obtained a very similar performance when reducing or augmenting the number of images used to train it. This suggests that the method is robust and that it could work well on new images.

The segmenter module, when compared to a manual segmentation performed by a clinician on 773 images, reached an average Jaccard index of 84.9%, with only 14.9% of the ROIs having less than 50% overlap with the experts’ ROIs.

### Clinical implications

The aforementioned results mean that the method would have detected almost 9 of 10 malignant cases and that almost every negative result was correct. Only a 2.2% of malignant cases would have been left undetected with a reasonable false positive rate, making the method ideally suited for high-risk patient screening. Its accuracy was comparable to the reported 75-84% accuracy of the clinician’s diagnostic when using dermoscopy after proper training^9,10^.

The use of this method could help clinicians during skin cancer monitoring and diagnosis and improve early detection. Used in a dermatology center to screen people at risk of developing skin cancer, it could help to derive quickly potential positive cases to the specialized dermatologist unit for further analysis. The method could help clinicians to better diagnose skin lesions avoiding the differences in terms of accuracy between countries, hospitals or even between clinicians of the same center (especially between a junior clinician and an experienced one). Finally, its automated nature should make it easy to install and use.

The proposed method will be integrated into an online platform (www.quantusskin.org), making it available to any professional. Other commercial tools are available in the market with similar purpose such as MelaFind®^28^ (a machine that gives the doctors more information about the lesions) or SkinVision^29^ (a mobile app that give the patient the risk of skin cancer from a photography of a lesion). However, none of them has a comparable detection rate to what reported in this study.

Naturally, we realize that the evaluation performed in this study, using online images does not constitute a clinical study. The results of this study need to be validated through a real prospective clinical study in a dermatologic center. This is the only way to observe the performance of the proposed methods on a real population.

## Strengths and limitations

This study has strengths and limitations. The main strength of this study is that the proposed method was trained and tested with a very large dataset of images (56,826) coming from 2 of the larger online databases worldwide (ISIC 2019 and ISIC 2020). This means that the images came from different centers and acquisition machines around the world so the data used is inherently multi-center and multiple equipment and operators were responsible for the acquisition of the images, making the study more robust and its results more reliable. Moreover, by combining the two datasets, the final prevalence (15.6%) is much more in accordance with a screening scenario of high-risk patients (13.6% reported in *Haenssle et al. 2016*^7^) compared to the use of ISIC 2019 alone (prevalence 33.4%), used by most prior works. Another strength is that the proposed method was designed striving for simplicity, using a single learning model, avoiding ensembles of models and other technical intricacies often used by many ISIC participants which can result in overfitting to data.

One of the limitations was that we were not involved in data acquisition and therefore we cannot be entirely sure of the correctness of the data used. However, these are public datasets widely used by the scientific community, so the assumption that data is correct is not entirely senseless. Another limitation is perhaps that we did not explore if demographic information (patient’s age, patient’s gender and lesion location) could be incorporated into the classifier to improve results. In this study we focused on developing a fully automated method and reporting the power of image analysis on its own. However, considering that the clinical variables were statistically correlated with respect to clinical outcome, this will be researched in future studies.

## Conclusion

To conclude, this study showed that a novel method based on Deep Learning could prove useful for the early detection of malignant skin lesions from dermoscopic images. Its superior accuracy compared to reported accuracy of dermatologists’ visual inspection and its high NPV show promise to be used during a first screening of patients, deriving the rest of patients to the specialized dermatologist unit for in-depth analysis and diagnosis. These results need to be validated through a prospective clinical study.

## Data Availability

All te data used in this study were downloaded online from the International Skin Imaging Collaboration (ISIC) Archive. The database used are composed of images provided from HAM10000 dataset, MSK dataset, BCN_20000 dataset and ISIC 2020 Challenge dataset. These datasets are licensed under a Creative Commons Attribution-NonCommercial 4.0 International License (CC-BY-NC).

https://challenge2020.isic-archive.com/

https://challenge2019.isic-archive.com/data.html

## Author contributions

D.C-G. was in charge of the study design and writing of the manuscript. He helped with the design of the method and with main experiments and performed all the statistical analysis of the results. He performed the literature review and wrote the first manuscript draft.

C.L. was in charge of data management and experiments. He helped to design the method, managed and downloaded all the data and performed the main experiments. He provided feedback on the manuscript.

X.P.B-A. was the study main supervisor. He helped with the design of the method and planned and supervised all experiments. He gave support with literature review and results analysis and wrote major sections of the manuscript.

## Data and materials availability

All data associated with this study is available in the main text and comes from public datasets. The developed AI method is available at www.quantusskin.org.

